# Deep Learning Based Long Term Mortality Prediction in the National Lung Screening Trial

**DOI:** 10.1101/2022.01.12.22269152

**Authors:** Yaozhi Lu, Shahab Aslani, Mark Emberton, Daniel C. Alexander, Joseph Jacob

**Affiliations:** Centre for Medical Image Computing, University College London, London WC1V 6LJ, UK; Department of Computer Science, University College London, London WC1E 6BT, UK; Department of Respiratory Medicine, University College London, London WC1E 6BT, UK; Division of Surgery and Interventional Science, University College London, London W1W 7TS, UK

**Keywords:** Computed Tomography, Deep Learning, Lung, Saliency Map

## Abstract

In this study, the long-term mortality in the National Lung Screening Trial (NLST) was investigated using a deep learning-based method. Binary classification of the non-lung-cancer mortality (i.e. cardiovascular and respiratory mortality) was performed using neural network models centered around a 3D-ResNet. The models were trained on a participant age, gender, and smoking history matched cohort. Utilising both the 3D CT scan and clinical information, the models can achieve an AUC of 0.73 which outperforms humans at cardiovascular mortality prediction. By interpreting the trained models with 3D saliency maps, we examined the features on the CT scans that correspond to the mortality signal. The saliency maps can potentially assist the clinicians’ and radiologists’ to identify regions of concern on the image that may indicate the need to adopt preventative healthcare management strategies to prolong the patients’ life expectancy.

## I. Introduction

### A. Overview

**C**ARDIAC and respiratory illnesses are the leading causes of mortality globally [1], [2], especially amongst older age groups. The ageing global population means that greater numbers of patients with multimorbid conditions are utilising healthcare services with increasing frequency and for ever more complex problems. Responding to such growing health-care needs requires cost-effective approaches for the early detection of disease. Early detection allows timely intervention before diseases become irreversible. In this study, a joint human-computer approach is proposed to identify imaging features on CT that are predictive of mortality in patients undergoing Lung Cancer Screening (LCS).

Annual Computed Tomography (CT) imaging in LCS studies has been demonstrated to be an effective screening tool for the early detection of lung cancer, reducing lung cancer mortality in the National Lung Screening Trial (NLST) [3], [4], [5]. In this study, CT scans from NLST are examined with a 3D-ResNet [6], [7] to predict imaging features associated with long-term mortality outcomes. To allow patients and clinicians to understand the morphological basis for the mortality signal on the CT image, it is imperative that the model is able explain the relationship between the clinical outcome data and the imaging labels. Saliency map methods [8], [9] are popular approaches to highlight relevant features on images that can explain deep learning models and localise contributing features to predictions. The visual interpretation of model classifications through saliency maps, for example when predicting long-term mortality, can aid radiologists in the identification of unsuspected pathology on a CT scan. In turn, this may allow early interventions that may prevent or delay the occurrence of adverse health events, thereby prolonging a patients’ life expectancy. This is of particular relevance to LCS where patients have numerous comorbidities but where detecting lung cancer is typically the overarching aim of the radiologist.

### B. Literature Review

Lung cancer and Cardiovascular Diseases (CVD) share several similar risk factors including smoking (both active and passive) and exposure to fine particulates from air pollution [10]. Though the pathophysiological mechanisms differ, it has been shown that smoking leads to increased mortality risk from both lung cancer and CVD [11], [10]. It is therefore logical that patient cohorts enriched with heavy smokers, such as LCS studies (e.g.the National Lung Screening Trial (NLST)) [3], [4], [5] can be used for the prediction of CVD-related mortality.

Recently, attempts have been made to predict CVD-related mortality in the NLST cohort [12], [13]. These studies have adopted Deep Learning (DL) based approaches to analyse NLST Low-Dose CT scans (LDCT). As demonstrated in van Velzen et al. 2019[12], a Convolutional Autoencoder (CAE) was trained to derive abstract image features and the features were fed into 3 separate classifiers to predict CVD-related mortality. The CAE encoded the automatically extracted 3D LDCT volume around the heart and exported the image features to the subsequent classifiers. The support vector machine classifier achieved performances in terms of Area under ROC curve (AUC) of 0.72. Though the study recognised the value of using clinical information for prediction, including hand-crafted variables such as the Coronary Artery Calcium (CAC) score [14], [15] which is a known predictor of CVD, such information was not utilised in making predictions. Instead, the study demonstrated that it is possible to predict CVD-related mortality from LDCT scans alone.

Predicting CVD-related mortality was further improved by Guo et al. 2020 [13]. A multimodal approach was adopted in this study where models incorporated both LDCT imaging information and handcrafted features to make mortality predictions. Firstly, for imaging data, a dual branch CNN network was adopted. Two 2D ResNets [6] were used to analyse manually selected 2D axial slides at 2 different resolutions/magnifications: the whole lung region and an automatically cropped cardiac region. For the clinical data, manually derived metrics such as CAC, were used to train a linear support vector machine classifier. When the contributions of the imaging features and clinical data were optimised, this approach improved the AUC performance to 0.82. Both approaches show improvement over human performance in this regard. In fact, as reported by Guo et al., visual inspection of the coronary artery calcium measure by a radiologist could only achieve performance with an AUC of 0.64. However, despite the improvement in classification performance, there are a few drawbacks that require further study. The 2D axial slices were chosen from the 3D LDCT volumes based on the visibility of the coronary artery and thus the task complexity for the CNNs was reduced considerably. Additionally, the amount of human effort in curating the imaging data means that the approach can not scale well to real-world clinical scenarios.

One way to lessen the burden on clinicians to provide image-derived variables to models would be to automate the CAC score acquisition. The deep-learning based approach proposed by van Velzen et al. 2020[16] demonstrated the feasibility of scoring CAC and thoracic aortic calcification (TAC) automatically. More importantly, it had been shown that the model can be performed on CT scans derived using a variety of different CT imaging protocols.

Other than the direct prediction of mortality, the NLST cohort has also been analysed for long-term mortality risk stratification. In Lu et al. 2019 [17], a 2D CNN (inception-v4 architecture) was used to analyse chest radiographs in the NLST cohort. The study used Gradient-weighted Class Activation Maps (GradCAM) [9] to isolate the contributing imaging features. As illustrated by the classification maps [17], the deep learning model tended to focus on the cardiac region to search for cardiovascular and respiratory mortality signals. The results demonstrate that the proposed approach can stratify long-term mortality risk, and help identify patients that might benefit the most from preventative interventions.

### C. Objectives

This study aims to use both 3D LDCT lung volumes and readily available clinical data from NLST to make long-term mortality predictions, with minimum human input. More importantly, we aim to use saliency maps to identify contributing features on CT images that link to long-term mortality, particularly non-lung-cancer-related mortality. This can help radiologists and clinicians diagnose pathology that might not be obvious at first glance, or help corroborate the presence of subtle but important damage. Such an approach, highlighting neglected features on CT imaging in lung cancer screening populations may speed up decision making for radiologists and clinicians and help optimise patient care.

The key contributions of the study are as follows:

- A 3D deep-learning-based approach to predict long-term outcome based on imaging of the thorax.
- Propose an approach to identify areas of concern on CT volumes that by allowing preventative interventions can improve healthcare efficiency and utilise personalised intervention to improve patient quality of life.

## II. Methodology

### A. The National Lung Screening Trial

The National Lung Screening Trial was a multi-centered lung cancer screening study conducted in the US from 2002 to 2007 [3], [4], [5]. 53,454 heavy smokers, aged 55-74 years, who were at high risk for developing lung cancer were recruited. They were randomly assigned to either the low-dose CT (LDCT) branch (26,772 participants) or the chest radiography branch (26,732 participants) of the study. Three annual screening attendances for CT imaging, denoted as T0-T2, were conducted. The NLST study’s primary endpoint was mortality. The participants’ survival status and the cause of death were ascertained through evaluation of death certificate ICD-10 (International Classification of Diseases, 10th edition) codes. In the LDCT branch, the main cause of death (as of the end of 2009) was cardiovascular disease (26.1%), followed by lung cancer (22.9%) and then other types of cancer (22.3%) [4]. In 2015, the survival status of the cohort was updated. In our study, the latest LDCT scan (i.e. T2 screening) and accompanying clinical data, were used to predict long-term mortality in the NLST population. The participant’s survival status in 2015 was chosen as the ground-truth label.

### B. Dataset Selection

Based on the patients’ mortality status in 2015, the NLST dataset can be grouped into 3 classes: lung-cancer-induced mortality (LC), non-lung-cancer-induced mortality (NL), and alive (AL). The first group (i.e. LC) was withheld for later use to evaluate the ability of saliency maps models to localise known imaging features associated with patient mortality. For patients in the second group, only cases where the cause of death related to cardiac (ICD-10 codes: I10-I52) or respiratory diseases (ICD-10 codes: J00-J99) were selected. Furthermore, only cases with all three screenings timepoints (i.e. T0, T1 and T2) and with CT scan thickness in the axial plane of no more than 2.5mm were kept. The former criterion aimed to avoid bias introduced by patients who left the trial early for unknown reasons.

Of the elligible NL cases (n=873) 180 cases were randomly selected for analysis. To ensure a homogeneous dataset, scans with streak artefacts in the cardiac region (n=12) and patients with severe forms of thoracic spinal scoliosis (n=2) were excluded, resulting in a study population of 166 cases labelled as dying of either cardiac or respiratory death. The selected NL mortality cases were age, gender, and smoking history matched in a 1:2 ratio with a control population of participants alive at the end of the 2015 follow up period (the AL class). The final study population therefore comprised 498 cases, a third of whom (n=166) had died of cardiac or respiratory causes and n=332 remained alive in 2015.

As a result of NLST’s inclusion criteria of having a prolonged smoking history (i.e. minimum pack-year of 30), the pack-year distribution in the study population is skewed to the right which is illustrated in Fig. 1. The non-lung-cancer mortality class (NL) was older and had a greater smoking history than the control class (AL). To allow equivalent matching, criteria were relaxed to have a tolerance of ±10 pack-year and ±5 year age difference. As illustrated in Fig. 1, the pack-year and age distribution of the control cases were shifted to the right to satisfy the matching criteria.

**Fig. 1:**
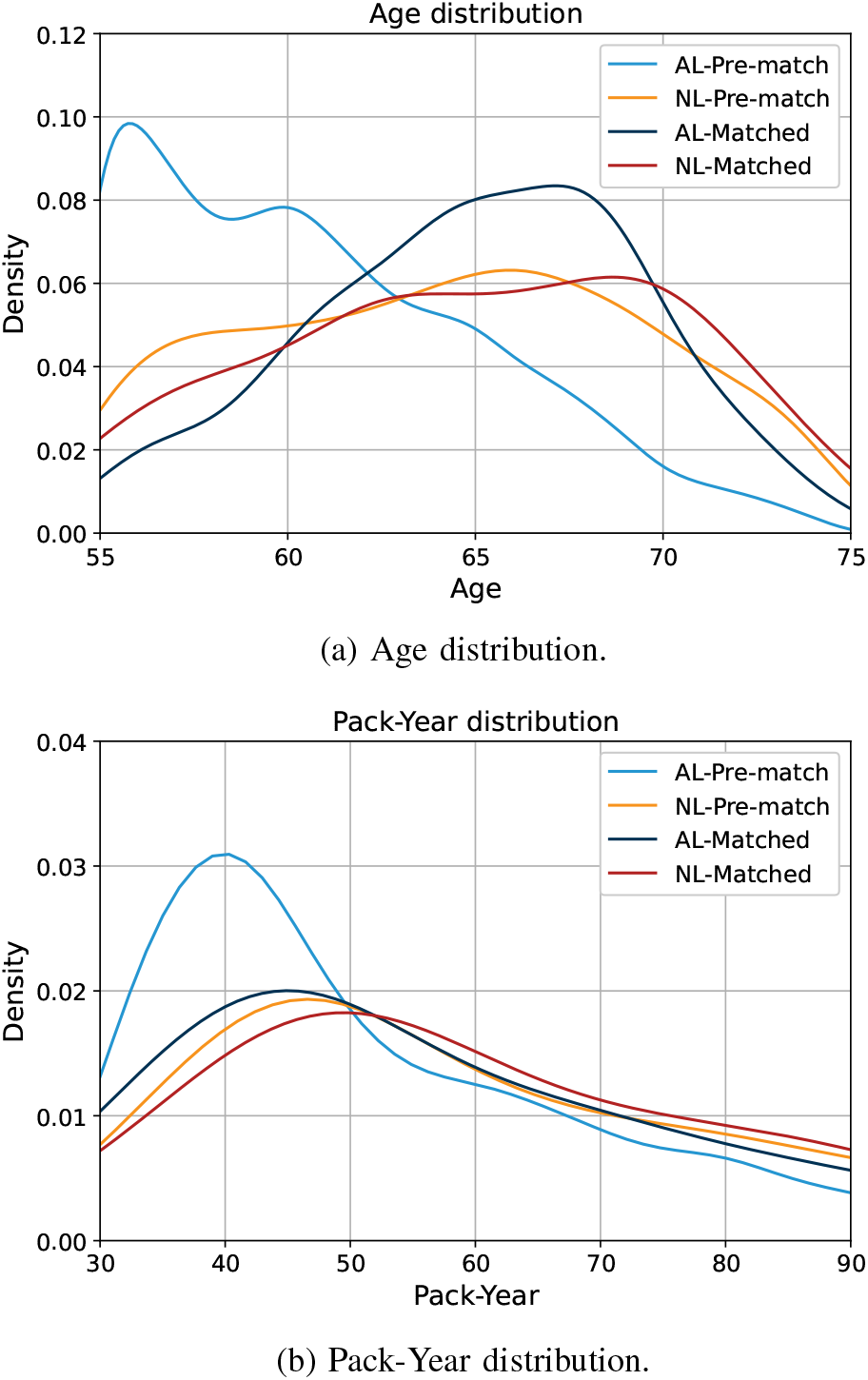
Pack-Year and age distribution of the matched dataset.

### C. Lung CT Volume Pre-Processing

The pre-processing approach used in Liao et al. [18] was adopted for this study. In essence, the lung masks were extracted from the raw DICOM image through convex-hull-based methods. The masks were then dilated and combined. The Hounsfield Unit (HU) range was clipped to an interval -1200 HU to 600 HU. The range was linearly normalised to the interval 0 and 255. The regions outside the masks corresponding to the surrounding tissue were filled with an average value of 170. The pre-processing pipeline was applied over all axial layers to extract the lungs in three dimensions.

Given that cardiovascular disease contributes to majority [4] of the mortality seen in the NLST dataset, it was felt important to preserve the cardiac region. Thus, an additional convex-hull calculation was performed on the joint lung masks to recover the CT information overlying the heart. The comparison between the original approach and the modified approach on the same axial slice is illustrated in Fig. 2. The region in blue corresponds to tissue outside the masks which was given an average intensity of 170. As is evident from Fig. 2(b), the modified pipeline preserves the cardiac region during pre-processing.

**Fig. 2:**
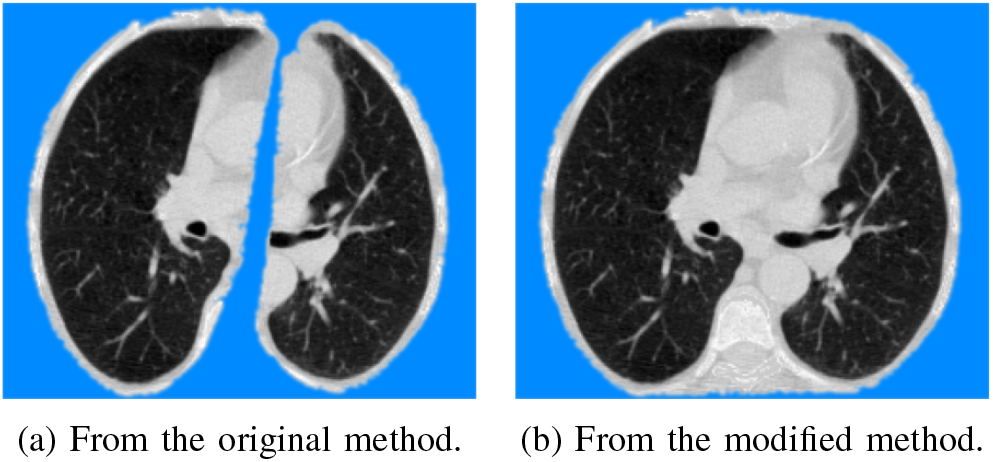
Comparison of the two pre-processing methods.

### D. Machine Learning Models

A two-tier approach was adopted to examine the effectiveness of different types of data, i.e. medical imaging and clinical data, in making mortality predictions. Firstly, non-deep-learning models were used to evaluate classification performance using only clinical data. Secondly, deep-learning-based methods were applied to the medical images to predict mortality. Finally, to assess whether the clinical information complements the CT scans in making mortality predictions, both clinical and imaging data were combined.

#### 1) Non-Deep-Learning Models

The Support Vector Machine (SVM) classifier (Model A), the Gradient Boosting Machine (GBM) classifier (Model B), and the Random Forest (RF) Classifier (Model C) were used to analyse the clinical data in the NLST dataset. The clinical data contained patient demographics, previous disease diagnosis, age, smoking history, and pack-year etc. A grid search with cross-validation was used to optimise the parameter settings for the three models. In addition to performing long-term mortality classification, the tree-based methods have the benefit of impurity-based feature selection. 11 features, tabulated in Table I, were selected to complement the CT imaging when making mortality predictions in the later part of the study. The 4 principal components encapsulating time information consistently ranked among the most important mortality-predicting feature for both tree-based models.

**TABLE I:** Selected clinical features.

| Feature | Description |
| --- | --- |
| BMI | Body mass index in $kg/m^2$ |
| gender | Patient's gender |
| diagchas | Pre-NLST diagnosis of childhood asthma |
| diagemph | Pre-NLST diagnosis of emphysema |
| diaghear | Pre-NLST diagnosis of heart diseases or heart attack |
| diaghype | Pre-NLST diagnosis of hypertension |
| invaslc | Lung-cancer related invasive procedure |
| PCA 1-4 | Principal components of the patient's smoking history.<br>The underlying features are:<br>smoking years, pack-year, age at smoke onset,<br>age at trial randomisation, and average number of<br>cigarettes per day |

#### 2) Deep-Learning

The deep-learning-based models in this study were based on a 3D implementation of the ResNet [6]. The pre-trained weights from Chen et. al.[7] were used for transfer learning purposes. The models were originally trained for medical imaging (CT images) segmentation tasks and had been shown effective in performing pulmonary nodule classification through transfer learning.

Two variants of the neural network were investigated: (1) the CT volume-only model (Model D), and (2) the multimodal model combining clinical and imaging data (Model E). They differ in the input data utilised and the corresponding ResNet-based architectures are illustrated in Fig. 3 and 4. The 3D ResNet backbone together with the pre-trained weights were used to analyse the input 3D CT volume. To transfer the pre-trained model to long-term mortality prediction in this study, the 3D ResNet backbone’s learning rate was reduced to 1/10 of the rest of the network. The output from the ResNet backbone was converted to a 1D tensor after passing through a global adaptive average pooling layer. In the CT-scan-only model (i.e. Fig. 3), the tensor was passed through 2 fully connected layers (including dropout with *p* = 0.5 and ReLU activation) with 512 and 32 neurons respectively. By contrast, in the multimodal version (i.e. Fig. 4), the clinical data (11 by 1 tensor) was concatenated with the output from the first fully connected layer before passing through the second layer with 43 neurons. Applying the concatenation at the later layer had the aim of providing more weight to the clinical information.

**Fig. 3:**
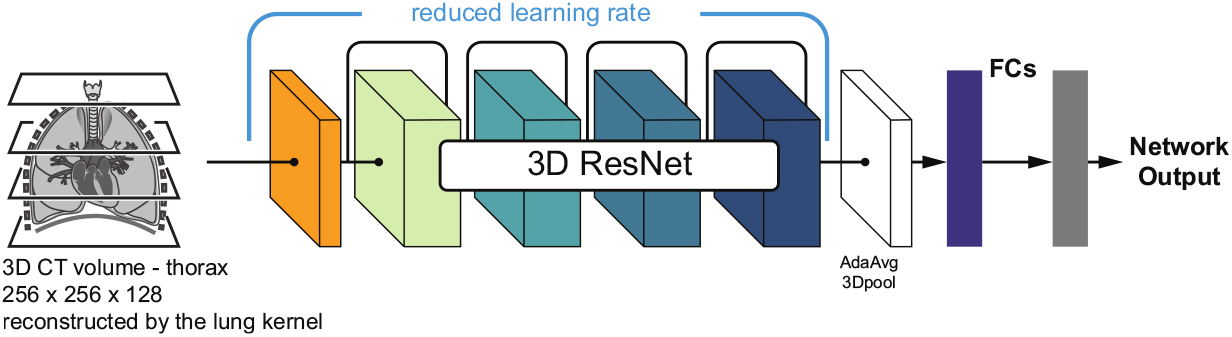
3D ResNet architecture for transfer learning (Model D). CT scans alone are passed to the network.

**Fig. 4:**
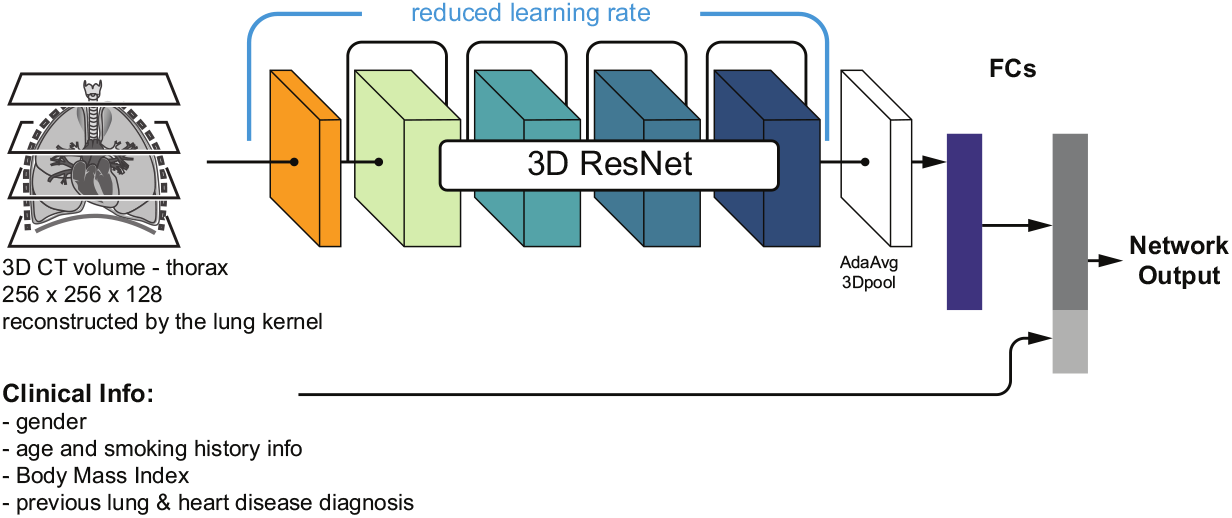
3D ResNet architecture for transfer learning (Model E). Both CT scans & clinical data are passed to the network.

To counter the class imbalance in the 1:2 matched dataset in this study, a weighted random sampler, which assigns a sampling probability inversely proportional to the class size was utilised. This approach attempts to produce, on average, balanced batches during training. Cross entropy loss was used as the loss function for the binary classification.

For the multimodal model as illustrated in Fig. 4, the two branches of the network were trained separately. The imaging branch of Model E inherited the weights from Model D and the training of the CNN portion was not optimised while training Model E. In contrast, the clinical data branch was optimised during training and the clinical measures were normalised to the range between 0 and 1. The alternative training strategy where the two branches, i.e. medical imaging and clinical data, are jointly trained were found to result in inferior mortality prediction performance.

### E. Saliency Maps

The saliency map methods were used to interpret the neural networks and to localise the contributing features to the predictions. As discussed earlier, performing saliency-map-based checks on the withheld lung cancer mortality class provided confidence in the trained models. In a clinical setting, saliency map visualisation of pathological regions on the CT contributing to mortality risk are crucial for clinician interpretation of potentially opaque neural network classifications. Saliency maps can also focus the attention of clinicians on potentially neglected or unrecognised structures on CT that are contributors to morbidity and mortality. This is particularly important in a setting where the detection of lung cancer typically takes primacy.

Common saliency map methods include the gradient approach [8], Gradient-weighted Class Activation Mapping (GradCAM) [9], Guided Backpropagation [19], and Guided GradCAM [9] etc. As elaborated in Adebayo et el. 2018 [20], the Guided Backpropagation and Guided GradCAM approaches tend to generate saliency maps that are independent of the data and model parameters, and cannot be relied upon to explain the model’s class prediction. The gradient approach and GradCAM are free from such shortcomings and are the approaches adopted in this study. Though both approaches can highlight relevant features for model predictions, the GradCAM approach tends to have lower resolution than the gradient approach, and thus can be limited when focusing on subtle/small features.

To filter out noise, thresholding was applied to the vector extracted from the last convolutional layer before it was extrapolated in 3D to the same dimension as the input CT volume. The rationale of applying thresholding before rather than after the interpolation step was to maximise the localisation capability of the resulting GradCAMs.

### F. Experiments

The 3D CT volumes were interpolated into a 256 by 256 by 128 volume before being passed into the neural networks. The 10-layer version of the 3D ResNet backbone was adopted in this study as it was felt that deeper models might only lead to marginal performance improvement. The Adam algorithm [21], which is an adaptive learning rate optimisation algorithm, was implemented to train the network with an initial learning rate of *1E-4*. The training was regularised by L2 regularisation (with weight decay parameter of *5E-3*). The networks were trained over 1,000 epochs.

To assess the performance of the models, 5-fold cross-validation was performed. Of the 498 cases in the study dataset, 48 were reserved for validation. The remaining 450 cases were used as training and testing sets and were split into 5 folds of 90. The relative class composition (i.e. 1:2 dead vs alive cases) was maintained in all the subsets. The selection of cases in the cross-validation folds was kept the same for all the models used. In each of the 5 cross-validation experiments, a machine-learning /deep-learning model was trained and evaluated.

To compare the performance with related studies [12], [13], the models were assessed with the Area Under the Curve (AUC) metric. The average value and the standard deviation of the AUC values from the 5-fold cross validation were used to gauge the overall performance of the models. Additionally, other performances metrics such as sensitivity, specificity, and F1 scores were also examined.

The non-deep-learning models (e.g. SVM) in this study were implemented through the Scikit-Learn library while the neural networks were implemented through the PyTorch library. Each neural network was trained with a batch size of 24 on a single Nvidia RTX 8000 GPU.

## III. Results

The median age in dataset used in this study was 66 and the median pack-year history was 54. Among the mortality cases, as shown in Fig. 5, the time difference between the latest screening point Ii.e. T2) and the time to patient death ranged from 0 to 10 years. Overall, 53.6% (89 patients) of the NL deaths were caused by cardiovascular diseases whilst the remainder (46.4%, 77 cases) were due to respiratory illnesses.

**Fig. 5:**
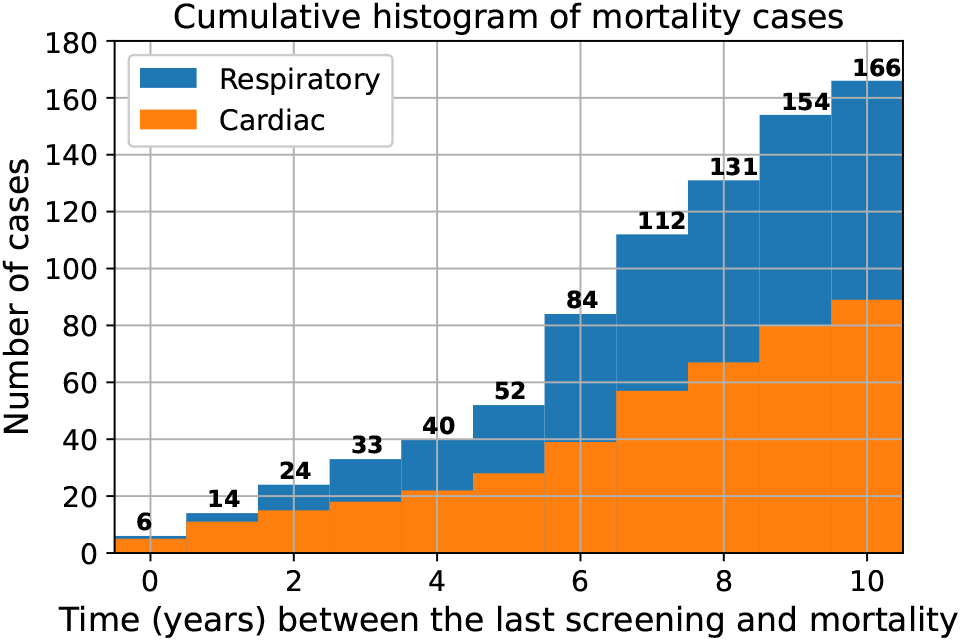
Cumulative distribution of the survival time.

The performance metrics of the models, in terms of the average performance of the 5-fold cross-validation, are tabulated in Table II. Models A-C, the non-deep-learning models were only trained with clinical data. Models D and E, the neural network models were trained on both medical imaging and clinical data. The corresponding ROC curves from the neural network models are grouped according to the type of input used and illustrated in Fig. 6. The composition of the cross-validation folds was maintained across the models.

**TABLE II:** Performance metrics.

| Model | Input | AUC (std) | Sensitivity | Specificity | F1 score |
| --- | --- | --- | --- | --- | --- |
| (A) Support Vector Machine | clinical data | 0.57 (0.05) | 0.66 | 0.48 | 0.49 |
| (B) Gradient Boosting Machine | clinical data | 0.58 (0.04) | 0.53 | 0.62 | 0.46 |
| (C) Random Forest | clinical data | 0.58 (0.03) | 0.84 | 0.33 | 0.53 |
| (D) 3D ResNet-10 | CT scan | 0.73 (0.03) | 0.71 | 0.68 | 0.61 |
| (E) 3D ResNet-10 | CT scan & clinical data | 0.73 (0.03) | 0.63 | 0.74 | 0.60 |

**Fig. 6:**
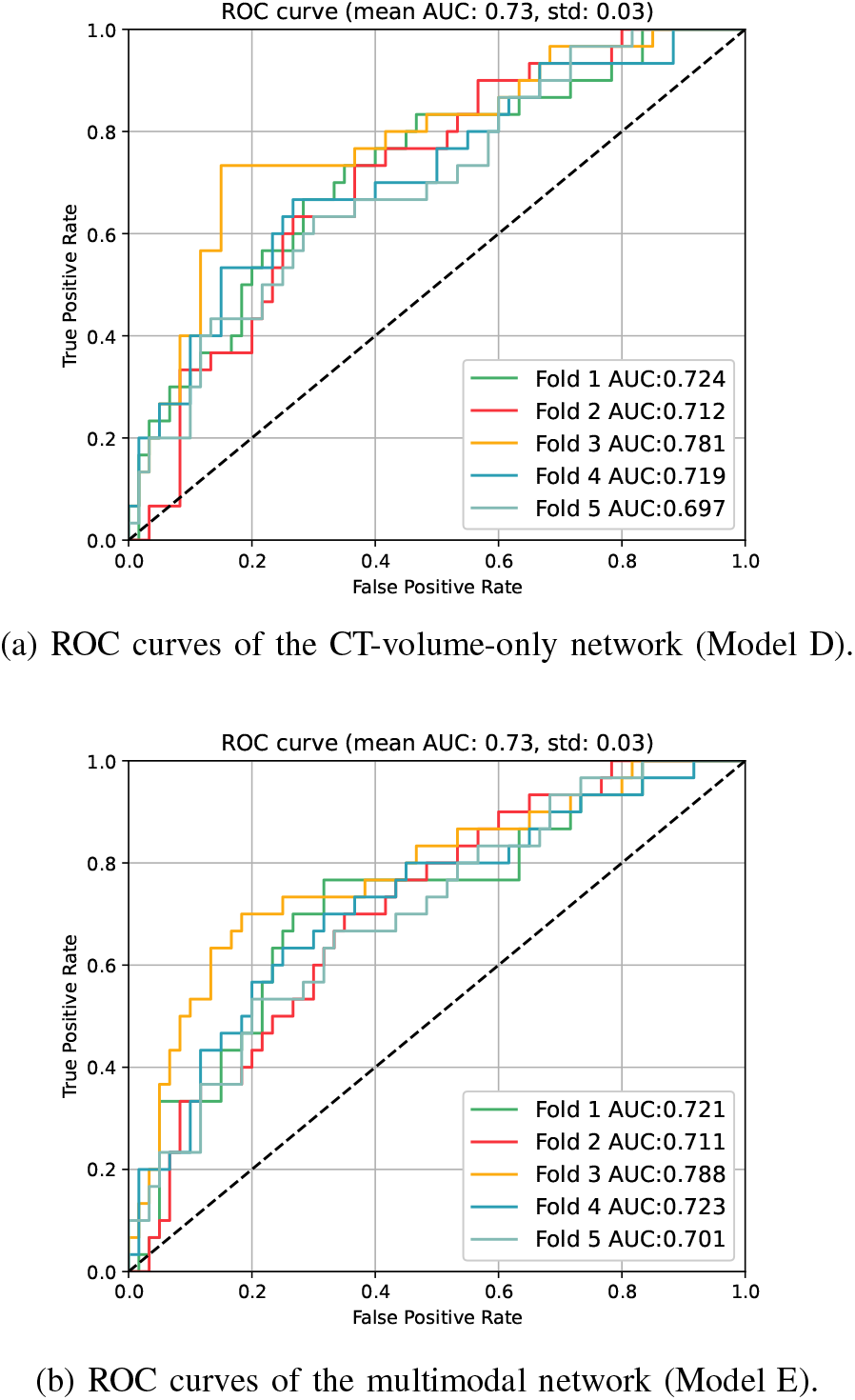
Performance comparison of the neural networks.

Among the non-deep-learning-based methods, the Random Forest model (Model C) achieved the best performance albeit limited with an average F1 score of 0.53 and an average AUC of 0.58. This suggests that the clinical data analysed in this study has limited utility in making long-term mortality predictions. The available clinical data was primarily collected to inform recruitment eligibility into the NLST and therefore might not have contained the most relevant prognostic variables. The deep-learning-based methods utilising medical images for classification performed better than non-deep-learning methods. The neural network model (Model D) trained with the 3D CT volume achieved a mean F1 of 0.61 and a mean AUC of 0.73.

## IV. Discussion

### A. Long Term Mortality Prediction

Though the clinical data denoting time information, i.e. PCA 1-4 in Table I, were ranked as the most important feature in mortality prediction by the tree-based models, their inclusion in Model E did not contribute to an improvement in performance over Model D. This was also true for clinical features encoding prior clinical diagnoses in patients. It is possible that the clinical variable of patient age, or more specifically biological age, might already be embedded in the imaging features, in the morphological appearances of the bones. Therefore, the introduction of chronological age through the clinical data offers limited additional information to the networks.

As shown in Table II, the performance of the current deep learning models for cardiovascular and respiratory mortality prediction, is on par with the cardiovascular mortality prediction models from van Velzen et al. 2019 [12] which has an average AUC of 0.72 and standard deviation of 0.07. Other performance metrics, i.e. sensitivity, specificity, and F1 score, were not available in the literature for comparison. While the benchmark study used an trained convolutional autoencoder to pass the extracted features to a SVM classifier, our models are trained in an end-to-end fashion. More importantly, the main motivation for identifying the respiratory mortality signal and visualising relevant areas on the CT image with saliency maps is to enhance clinical interpretability and confidence in our deep learning model predictions.

More recently, the cardiovascular mortality prediction study from Guo et al. [13] where a dual-ResNet was trained on manually selected 2D CT slices achieved a mean AUC of 0.76 and standard deviation of 0.10. When outputs from a SVM trained on handcrafted measures such as the CAC score, were tuned with the output from the dual-ResNet, the model achieved a mean AUC of 0.82. The performance improvement brought about by the handcrafted measures in Guo et al. [13], highlights the benefits of combining imaging features with known CVD mortality predictors. Therefore, for future work, we aim to develop an automated approach to derive these markers from the CT scans and use them in the multimodal prediction approach (i.e. Model E). In contrast to models from Guo et al. [13], the models in this study adopt an automated approach and thus lessens the burden on radiologists who would otherwise have to select CT slices of interest manually.

### B. Confirmation Using Saliency Maps

To evaluate the trustworthiness of the trained models, the saliency maps of the trained models were evaluated on previously held out lung cancer mortality cases using the gradient approach [8] and GradCAM [9]. Given that in these patients it was the lung cancer that accounted for patient mortality, the heatmaps, if performing appropriately would be expected to highlight the malignant nodules in the lung. 3 cases are illustrated in Fig. 7. The original inputs to the network, together with their saliency map overlays, are presented. The heatmaps are colored such that red denotes high activation suggesting the local imaging features contribute more to the network’s prediction. From the heatmaps it can be seen that the network can identify the cancerous nodules in the lung and are using these features appropriately when making mortality predictions.

**Fig. 7:**
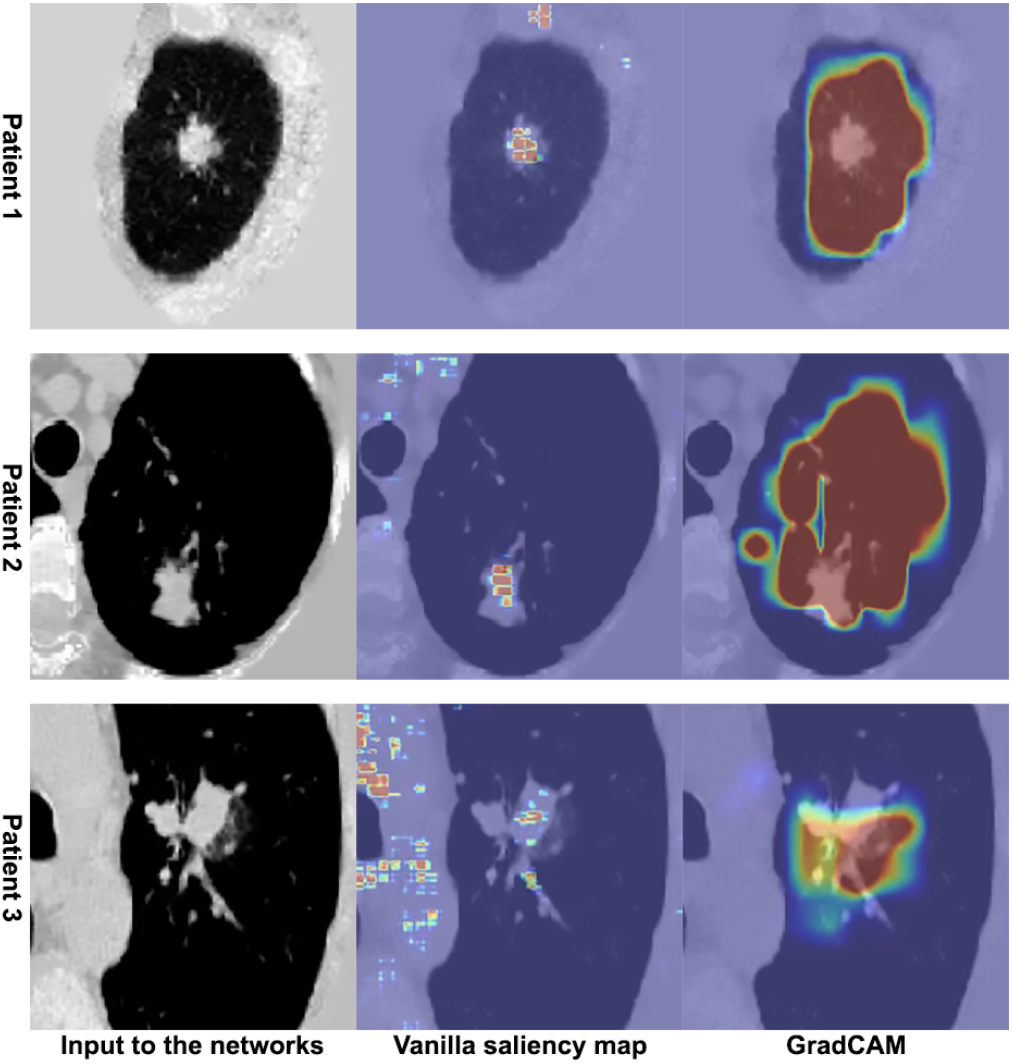
Sanity check (in axial view) on the saliency maps in CT volumes with cancerous lesion.

However, the current approach is not without potential limitations. Fig. 8 illustrates the GradCAM approach where the model has highlighted the malignant lesion in the right lung. The model has also highlighted the vertebra and the rib cage, which might intuitively have no direct link to cardiac and pulmonary-related mortality. Yet bone mineral density can act as a good indicator of patient health, particularly in patients with chronic lung diseases such as chronic obstructive pulmonary disease [22], [23]. Furthermore, various studies [24], [25] have demonstrated an association between low bone material density and coronary artery calcification. Accordingly, the signal of low bone material density shown using the GradCam approach may represent a relevant surrogate signal indicating both cardiovascular and respiratory-related mortality.

**Fig. 8:**
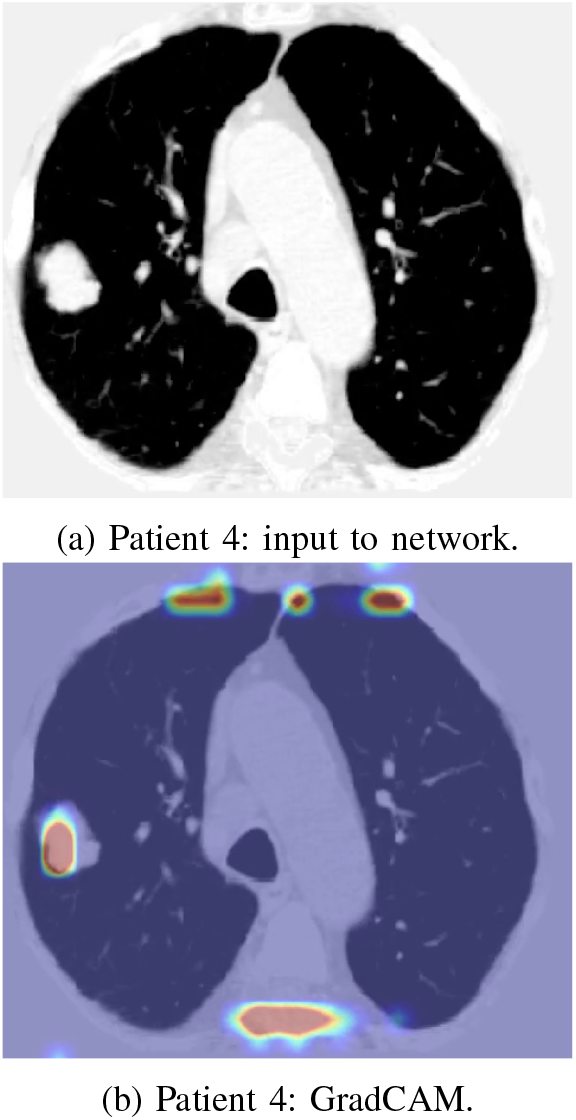
Saliency map with mixed signal.

## V. Conclusion and Future Works

The results shown in our study lead us to the following conclusions and directions for future studies.

1. The current method shows reasonable performance in predicting long-term mortality in the NLST dataset, illustrating the feasibility of performing mortality predictions from 3D CT scans, without handcrafted features.
2. The use of saliency maps shows promise as an aid for clinicians’ and radiologists’ in identifying neglected regions of the CT that might associate with mortality. This approach may facilitate the planning of personalised preventative interventions.
3. Given that cardiovascular diseases contribute to a significant portion of the non-lung-cancer mortality in the NLST dataset, it is reasonable to hypothesise that model performance can be improved by providing additional information from the cardiac region. One way to achieve this is by combining the mediastinal-kernel reconstructed cardiac CT volume with the lung-kernel-reconstructed lung volume used in this study. Accordingly, as a next step, we aim to develop a dual branch network for the imaging data. Additionally, we aim to explore an automated approach to calculate handcrafted measures, such as the CAC, to complement the automated image analysis in making mortality predictions.
4. In this study, the chronologically most recent scan (i.e. T2) in the NLST dataset was used to predict long-term mortality. The inclusion of the two earlier annual scans may add value by providing information on the progression of diseased sites in the lung. Such information can help identify patients who have more rapidly progressive diseases who would benefit more from early preventative interventions. To do so, we aim to explore longitudinal lung CT registration using all three screening time point CTs (i.e. T0, T1, and T2).
5. In addition to predicting long-term mortality outcomes, it would be worth exploring the quantification of damage in structures shown to be important on the saliency maps. Analysing quantitative variables in Cox Regression models to predict survival time may be an alternative way of identifying interpretable prognostic imaging biomarkers in LCS participants to facilitate risk stratification and personalised management in high-risk populations.

## Data Availability

All data produced in the present work are contained in the manuscript.

## Acknowledgment

This work was supported by the International Alliance for Cancer Early Detection, an alliance between Cancer Research UK [C23017/A27935], Canary Center at Stanford University, the University of Cambridge, OHSU Knight Cancer Institute, University College London and the University of Manchester.

The authors thank the National Cancer Institute for access to NCI’s data collected by the National Lung Screening Trial (NLST). The statements contained herein are solely those of the authors and do not represent or imply concurrence or endoresement by NCI.

## Notes

### Competing Interest Statement

The authors have declared no competing interest.

### Author Declarations

This study involves only openly available human data, which can be obtained from the National Lung Screening Trial (https://www.cancer.gov/types/lung/research/nlst).

